# Promoting Community Living and Participation Outcomes Among Young Autistic Adults: Promising Results for an Autistic-Delivered Peer Support Intervention

**DOI:** 10.64898/2026.10.18.26365838

**Authors:** Wei Song, Annemarie Clarke, Gregory Bannett, Katy Kaplan, Jamie Reese, Jonas Ventimiglia, Stacy Nonnemacher, Lindsay Shea, Mark S. Salzer

## Abstract

Autistic young adults experience substantial gaps in services supporting community living and participation. The Community Autism Peer Support (CAPS) program is a community-based intervention in which trained autistic peer specialists provide individualized support to autistic young adults pursuing self-determined goals related to community participation and independent living. We conducted a single-site, two-arm randomized controlled trial to evaluate the feasibility of implementing the CAPS program. Forty autistic adults aged 18–30 years were randomized to CAPS (n = 20) or services as usual alone (n = 20) and assessed at baseline, 3 months, and 12 months. Study findings showed that CAPS attracted autistic young adults with diverse demographic and diagnostic backgrounds, and most intervention participants remained engaged in services for extended periods. While the study was underpowered to detect statistical differences in outcomes because it was not a main focus of the research, a consistent pattern of results favored the CAPS condition across all outcomes. The largest positive signals were observed for overall and social quality of life, with additional favorable changes in internalized stigma, unmet needs, social functioning, loneliness, and positive autistic identity. Findings provide support for the feasibility and promise of an autistic-delivered peer specialist model focused on community living and participation. The consistent pattern of favorable changes supports continued development of CAPS as a person-centered approach to addressing outcomes important to autistic adults.

## Introduction

Autistic adults experience substantial gaps in opportunities related to employment, education, social relationships, and overall community participation.^1–4^ They participate in their community half as often as adults in the general population and are much less satisfied with their participation across social, civic, and recreational domains.^5^ These challenges are compounded by the loss of services after exiting the school system and smaller support networks.^6–8^ This period is particularly consequential, as young adulthood is a unique developmental period when individuals begin to seek autonomy and actively choose social roles (e.g., friendships, education, professions, leisure interests) that shape long-term quality of life.^9^ Greater engagement in meaningful social and community activities contributes to improved quality of life and reduces social isolation, anxiety, and stress for autistic individuals, but it can be challenging without support.^10–14^

Current services and interventions largely prioritize early interventions for autistic children and youth ^15,16^ and those that exist for adults primarily focus on social skills and employment, with limited effectiveness in both areas.^17–19^ While services aimed at enhancing employment outcomes are very important, autistic adults have an interest in a broad range of community participation domains and desire greater participation in other areas.^5,20,21^ For example, Song et al.^5^ found that more than half of autistic adults thought volunteering and going to a park or recreation center were important to them, but only a quarter of them participated in the past month. Effective services that target broad community participation during early adulthood could be essential for long-term functioning, mental health, and wellness.

Scholars have increasingly emphasized the involvement of autistic individuals as experts-by-experience in the design, delivery, and evaluation of supports and interventions. ^22,23^ This shift aligns with neurodiversity-oriented approaches that prioritize autistic perspectives, self-determination, and the development of supports that are responsive to the priorities and lived experiences of autistic people.^23,24^ Within this context, autistic-delivered peer support has emerged as an approach where there is a great deal of enthusiasm. Peer support is grounded in shared lived experience and emphasizes mutual understanding, empowerment, and practical support that may not be accessible through professional expertise alone.^25^ Emerging evidence suggests that autistic adults value opportunities to connect with autistic peers who share similar experiences and provide understanding, validation, and guidance related to navigating community life, relationships, services, and personal goals.^24,26^ A systematic review of 12 peer-support programs for autistic individuals indicated that they might improve self-esteem, academic performance, empowerment, and overall well-being.^24^ However, these programs primarily involved segregated group activities focused on leisure and interactions among program participants rather than emphasizing the pursuit of self-determined, independent living and community participation goals.

The Community Autism Peer Support (CAPS) program is a novel, autistic-delivered peer support intervention for autistic young adults that was developed through a partnership among autistic adults, family members, peer support professionals, service providers, researchers, and policymakers. CAPS involves trained autistic peer specialists who work one-on-one with autistic young adults to identify and pursue self-determined goals related to employment, education, relationships, leisure activities, and other life areas. A quasi-experimental evaluation of CAPS showed that the program was feasible and acceptable to both participants and peer specialists. ^27,28^ Autistic participants were highly engaged in the program, developed an effective working alliance with the autistic peer specialists, and were highly satisfied with the program. ^27^ Participants at the 3-month follow-up reported fewer unmet needs for community living and improved social functioning.^28^ Participants also reported increased engagement in other healthcare services (e.g., mental health services) and higher perceived support from other service providers.^28^ The absence of a control group and the short follow-up period, however, limit conclusions regarding the program’s effectiveness.

The aims of the current study were to refine the CAPS intervention, including the creation of a manual, and then determine the feasibility of implementing the CAPS intervention as part of a randomized, controlled trial (RCT) which involved assessing the likelihood of recruiting and enrolling adequate samples for a larger RCT while also testing data collection procedures. The sample size for enrollment (i.e., 40), was determined based on pragmatic considerations related to this objective instead of statistical power. Instead, the primary goal was to assess whether a future, fully powered RCT was likely to be successfully carried out. ^29,30^ While statistical power was limited, results from such studies can help determine whether there are any indications, or signals, of possible treatment effects, if interpreted with caution.^31,32^ Therefore, this study examines whether there are any indications that the CAPS intervention demonstrates possible promising effects, despite being underpowered, by examining outcome pattern differences between the conditions. The study is important because of relative novelty of autistic-delivered services and focus on promoting community living and participation using a rigorous research design.

## Emerging Practice

### Development of the CAPS Program

CAPS was developed to address the limited availability of community-based supports for autistic individuals transitioning to adulthood. It was designed as an autistic-delivered, peer support intervention where staff provide individualized, one-on-one support to other autistic individuals identifying and pursuing self-determined goals. The original 75-hour CAPS training program was developed by Community Behavioral Health (CBH), the Medicaid payer in Philadelphia, in collaboration with autistic self-advocates with significant experience as behavioral health providers and was overseen by a CAPS Advisory Board, consisting of self-advocates, caregivers, researchers, and professionals.

Building upon this foundation, our project refined the training and services by developing a manualized intervention focused on supporting participants in pursuing self-determined goals related to community participation. Although participation goals varied across individuals, common domains included employment, education, leisure and recreation, civic engagement, faith activities, social activities, parenting, and other valued community roles. Consistent with a holistic approach to community participation, the intervention also addressed individual and environmental factors that may facilitate or hinder goal attainment, including self-efficacy, self-advocacy, internalized stigma, coping, problem-solving, emotional regulation, access to services and community resources, and relationships with family members and friends.

The CAPS program was informed by several complementary peer support theories,^34^ including social comparison theory, ^35^ social learning theory,^36^ social support theory,^37^ and the valuing of experiential knowledge. ^38^ Together, these theories suggest that shared lived experiences allow autistic peer specialists to provide role modeling, emotional validation, practical guidance, and unique experiential knowledge in ways that promote hope, empowerment, self-efficacy, and positive identity.^34^ Unlike traditional therapeutic relationships, CAPS emphases mutual learning, participant choice, and strengths-based support, positioning autistic adults as experts in their own lives and as active partners in the support process.

### Description of the CAPS Program

The CAPS program involved four interrelated components: (1) peer support and mutual learning, (2) needs assessment, person-centered goal setting, and progress monitoring, (3) capacity building through lived experience, and (4) resource navigation and community connection. Peer support and mutual learning emphasized developing trusting relationships through shared lived experience, active listening, validation, encouragement, and mutual learning. Needs assessment, person-centered goal setting, and progress monitoring focused on identifying participants’ strengths, needs, and priorities, collaboratively establishing individualized goals and action plans, and regularly reviewing progress toward those goals.

Capacity building through lived experience involved sharing practical knowledge and personal experiences, developing coping, problem-solving, self-advocacy, and emotional regulation strategies, and addressing barriers to community participation. Resource navigation and community connection focused on connecting participants with formal and informal supports, facilitating access to services and accommodations, strengthening relationships with family and friends, and increasing engagement in community resources and activities. Table 1 summarizes the intervention components and the outcome areas they were hypothesized to impact.

**Table 1.** CAPS program outcomes connected to intervention strategies.

| Intervention Components | Hypothesized Outcomes |
| --- | --- |
| • Peer support and mutual learning;<br>• Capacity building through experience sharing; | Reduced internalized stigma |
| • Needs assessment, person-centered goal setting, and progress monitoring;<br>• Capacity building through experience sharing;<br>• Resource navigation and community connection; | Reduced unmet needs |
| • Needs assessment, person-centered goal setting, and progress monitoring;<br>• Capacity building through experience sharing; | Increased self-efficacy |
| • Capacity building through experience sharing; | Social functioning |
| • Peer support and mutual learning;<br>• Capacity building through experience sharing; | Increased positive identity; |
| • Peer support and mutual learning;<br>• Resource navigation and community connection; | Decreased isolation/loneliness |
| • Peer support and mutual learning;<br>• Needs assessment, person-centered goal setting, and progress monitoring;<br>• Capacity building through experience sharing;<br>• Resource navigation and community connection; | Increased quality of life |

Participants selected their community participation goals and documented their strengths and existing support related to their goals. The peer specialists then worked with participants to develop individualized support plans that included specific objectives, action steps, and strategies for addressing barriers. Ongoing meetings focused on reviewing progress, revising goals and action plans as needed, and providing support tailored to participants’ evolving needs. The CAPS services were designed to be flexible and individualized. While peer specialists and participants were encouraged to meet at least weekly, the frequency, duration, and format of contacts were determined collaboratively based on participants’ goals, preferences, and support needs. In the one-on-one sessions, peer specialists facilitated participation through peer support and mutual learning, participant-directed goal planning, capacity building, and navigation of community resources.

The CAPS program was implemented by an agency that offers extensive autism and peer support services. Two autistic peer specialists were recruited from among those who had completed a 75-hour peer specialist training program and were employed to deliver the intervention. The peer specialists then obtained additional training provided by the research team, including experienced autistic peer specialists, which involved a 10-hour refresher training covering the essential components of the peer specialist training and 12-hour training on the CAPS program model. Peer specialists received ongoing supervision and technical support throughout the intervention, which also included an autistic peer specialist who had significant experience providing similar supports.

## Evaluation Methods

### Study Design

A single-site, two-arm RCT was conducted in the city of Philadelphia between June 2023 and August 2025 (ClinicalTrials.gov ID: NCT05702658). A CONSORT flow diagram of the study procedure is outlined in Figure 1. The observation period was 12 months with assessments at baseline, 3 months, and 12 months. Data collection was conducted by trained research staff with experience with the population.

**Figure 1.**
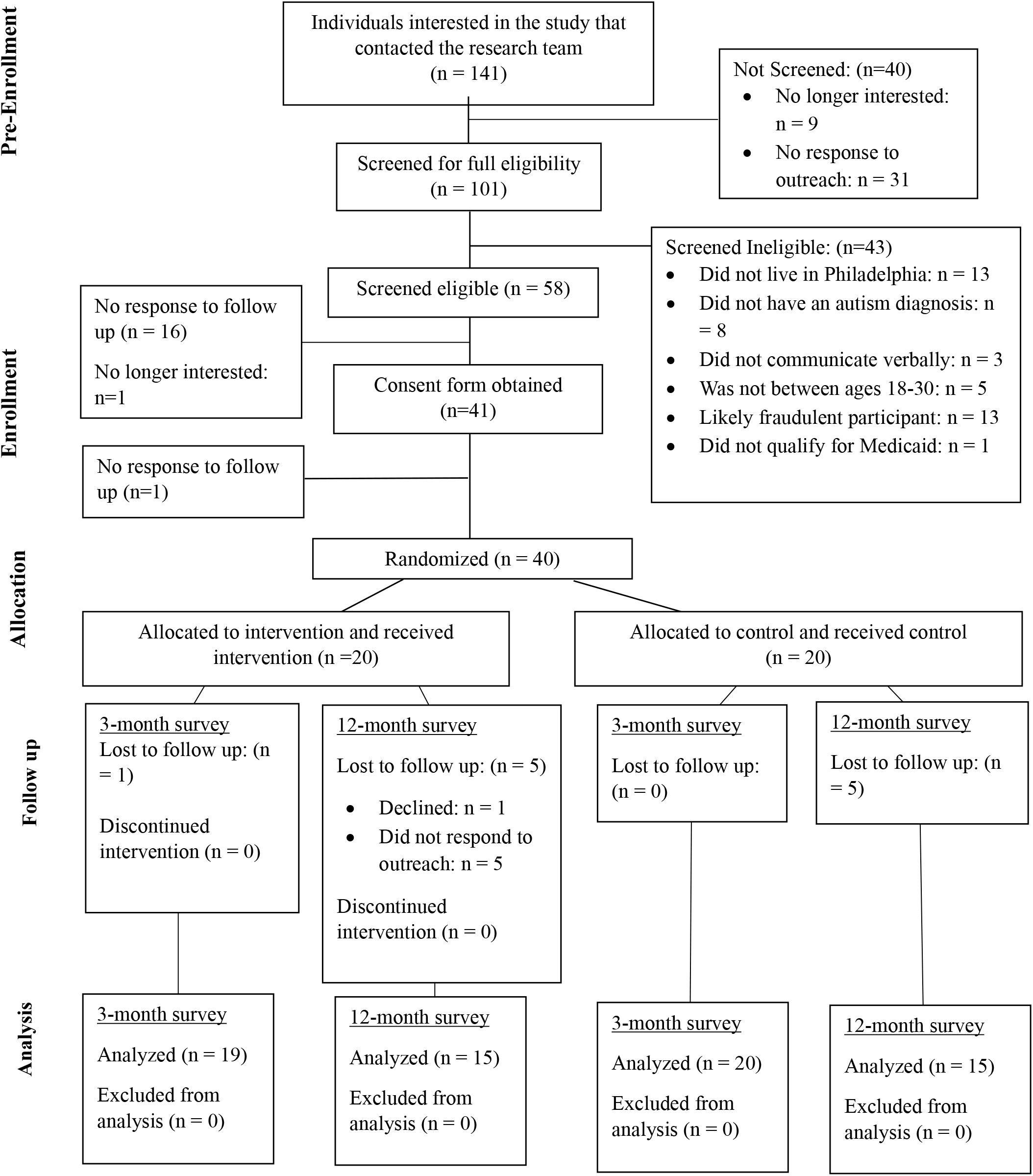
Study participant flow diagram.

### Participants

Participants met the following inclusion criteria: (1) 18-30 years old; (2) had a diagnosis of ASD from a licensed healthcare provider; (3) reported challenges in one or more major life areas (i.e., social, school, work, self-care); (4) able to communicate in written or spoken English; (5) able to provide informed consent, (6) lived in the city of Philadelphia, and (7) enrolled in or eligible for Medicaid. Participants were excluded if they were currently receiving peer support services elsewhere.

Participants were recruited through local autism service agencies, support and community groups, and local universities between April 2023 and August 2024. Again, the primary goal was to assess the feasibility of carrying out a large-scale RCT. One important finding related to this is that a total of 141 individuals contacted the research team with interest in participating and 101 individuals were screened to see if they were eligible. Forty-three out of 101 screened ineligible due to location, autism diagnosis, communication needs, age, and others (see Figure 1). Among the 58 eligible individuals, 18 did not respond to follow-up requests to meet. The remaining 40 individuals were included in the analysis.

### Procedures

Upon obtaining informed consent, research staff administered the baseline measures, performed the random group assignment and notified enrolled participants if they were randomly assigned to the intervention (=20) or the control/services as usual (n=20). The research staff were blind to the random allocation sequence and entered participants into the randomized service with permuted blocks of four. Follow-up interviews were conducted at 3- and 12-months post-baseline.

Participants in the intervention group received the CAPS intervention, described above, in addition to any healthcare, educational, or other services they were receiving. Participants in the service-as-usual condition also continued to receive whatever services they were already receiving but did not receive CAPS services. The overall attrition rate at follow-up time points was 2.5% (data obtained from 39 out of 40 participants) at the 3-month and 20% (32 of 40) at the 12-month timepoints. Ethical approval was granted by Drexel University IRB and City of Philadelphia IRB.

### Outcome Measures

#### Internalized Stigma

Two items adapted from the Brief version of the Internalized Stigma of Mental Illness (ISMI-10) scale ^39^ were used to assess internalized stigma. To increase relevance for autistic adults, references to “mental illness” were replaced with “autism.” The items assessed participants’ endorsement of negative beliefs about their value and ability to lead a fulfilling life as an autistic person (e.g., “I can’t contribute anything to society because I have autism” and “I can have a good, fulfilling life despite being an autistic person” [reverse scored]). Responses were rated on a 4-point Likert scale (1= “Strongly Disagree” to 4=“Strongly Agree”). Scores were calculated by averaging the two items, with higher scores indicating greater internalized stigma.

#### Self-Efficacy

The General Self-Efficacy Scale (GSES) is a 10-item scale^40^ designed to measure an individual’s perceptions regarding their ability to manage novel situations.

Participants were asked to rate their level of agreement with statements, on a 4-point Likert scale ranging from “Not at all true” (1) to “Exactly true” (4). Sample items include, “I can always manage to solve difficult problems if I try hard enough” and “Thanks to my resourcefulness, I know how to handle unforeseen situations.” Higher total scores represented greater self-efficacy.

#### Unmet Needs

A modified version of the Camberwell Assessment of Needs ^41^ was used to assess unmet needs in 23 life domains, including housing, taking care of one’s home, autism information and treatment, psychological distress, social life, intimate relationships, and other areas. It is a validated and reliable instrument that captures the total number of unmet needs, reported by each individual, that would support them in to living a healthy and fulfilled life.

#### Social Responsiveness

The Social Responsiveness Scale, 2nd edition (SRS-2) is among the most well-validated and frequently used measures of ASD symptomatology ^42,43^. It is a 65-item instrument with subscales in the domains of social awareness, social cognition, social communication, social motivation, and restricted interests and repetitive behavior. The SRS-2 Adult Self-Report Form was slightly modified to reflect extended age ranges and associated respondent characteristics ^44^. Raw scores were transformed to T-scores, which were used in the analysis ^44^.

#### Positive identity

The Disability Pride and Exclusion/Dissatisfaction subscales of the Questionnaire on Disability Identity and Opportunity (QDIO)^45^ were used to assess positive identity. QDIO is a measure developed to assess disability identity and orientations toward disabilities or conditions. In our study, the word “disability” was substituted by the word “autism/autistic” as appropriate. The disability pride (i.e., autism pride) subscale reflects perceived importance of or pride in autism being part of oneself (e.g., “Autism is an important part of who I am”). The exclusion/dissatisfaction subscale captured feelings of rejection or dissatisfaction with being autistic. Participants scored each item on a five-point Likert scale (1= “Strongly disagree” to 5= “Strongly agree”), with higher scores indicating more positive identity of autism.

#### Loneliness

Loneliness was assessed using the 3-item UCLA Loneliness Scale, a brief measure derived from the Revised UCLA Loneliness Scale ^46^. Participants were asked how often they felt that they lacked companionship, felt left out, and felt isolated from others. Items were rated on a 3-point scale ranging from 1 (“I often feel this way”) to 4 (“I never feel this way”). Total scores range from 3 to 12, with higher scores indicating less loneliness.

#### Quality of Life

Quality of life was assessed using items from the World Health Organization Quality of Life Instrument (WHOQOL-BREF; The WHOQOL Group, 1998) at the baseline and 12-month. Overall quality of life was measured using a single question (“How would you rate your quality of life?”) rated on a 5-point Likert scale from “Very Poor” (1) to “Very Good” (5).

Social QoL was assessed using the Social Relationships domain, which evaluates satisfaction with personal relationships, social support, and intimate relationships. Items were rated on 5-point scale from “Very Dissatisfied” (1) to “Very Satisfied” (5). The total domain score of social QoL was transformed to a 0-100 scale^47^, where higher scores indicated better social QoL.

### Statistical Analysis

First, all descriptive statistics for socio-demographic characteristics and outcome data were generated using R. Based on the intention-to-treat principle, we analyzed the data according to participants’ randomized assignment. All available outcome data were included in the longitudinal models. Generalized linear mixed models (GLMMs) were conducted to examine the changes in mean scores in individual outcomes. Fixed effects included study groups, time, and the group x time interaction. The primary interest was the group X time interaction, which estimates differential change in outcomes between study groups over time. Unstructured covariance structure was selected based on model convergence and fit indices. Degrees of freedom were estimated using the Kenward-Roger method to provide small-sample correction of standard errors. Models were estimated using restricted maximum likelihood (REML) in SAS.

As mentioned earlier, the study was underpowered, yet such studies can provide insights about possible treatment effects in large-scale effectiveness trials.^31,32^ To detect possible indications of CAPS effectiveness, within-group effect sizes on each outcome measure from baseline to 12 months were also calculated for both groups and then compared using Cohen’s D. Patterns in the direction of differences were also examined. For example, if positive changes favoring the CAPS condition were found on 75% of the outcome measures, this might be used as an additional indicator of potential effectiveness favoring CAPS.

## Results and Lessons Learned

### A Broad-Range of Autistic People are Interested in CAPS

CAPS attracted autistic young adults with diverse demographic and clinical backgrounds (Table 2). The mean age was 22.08 (SD=3.23). Participants ranged in age from 18 to 30 years (M = 22.08, SD = 3.23) and represented diverse racial and ethnic backgrounds, including 45.0% (n = 18) Black/African American, 37.5% (n = 15) White, and 17.5% (n = 7) identifying with another race or multiple races. Most participants identified as male (n = 29, 72.5%) and non-Hispanic/Latinx (n = 32, 80.0%). Although most participants had received an autism diagnosis during childhood, seven (17.5%) were diagnosed during adulthood, suggesting that the program was of interest to autistic adults with varied diagnostic experiences. The small sample size prevented the meaningful examination of potential group differences, but the groups appear comparable.

**Table 2.**
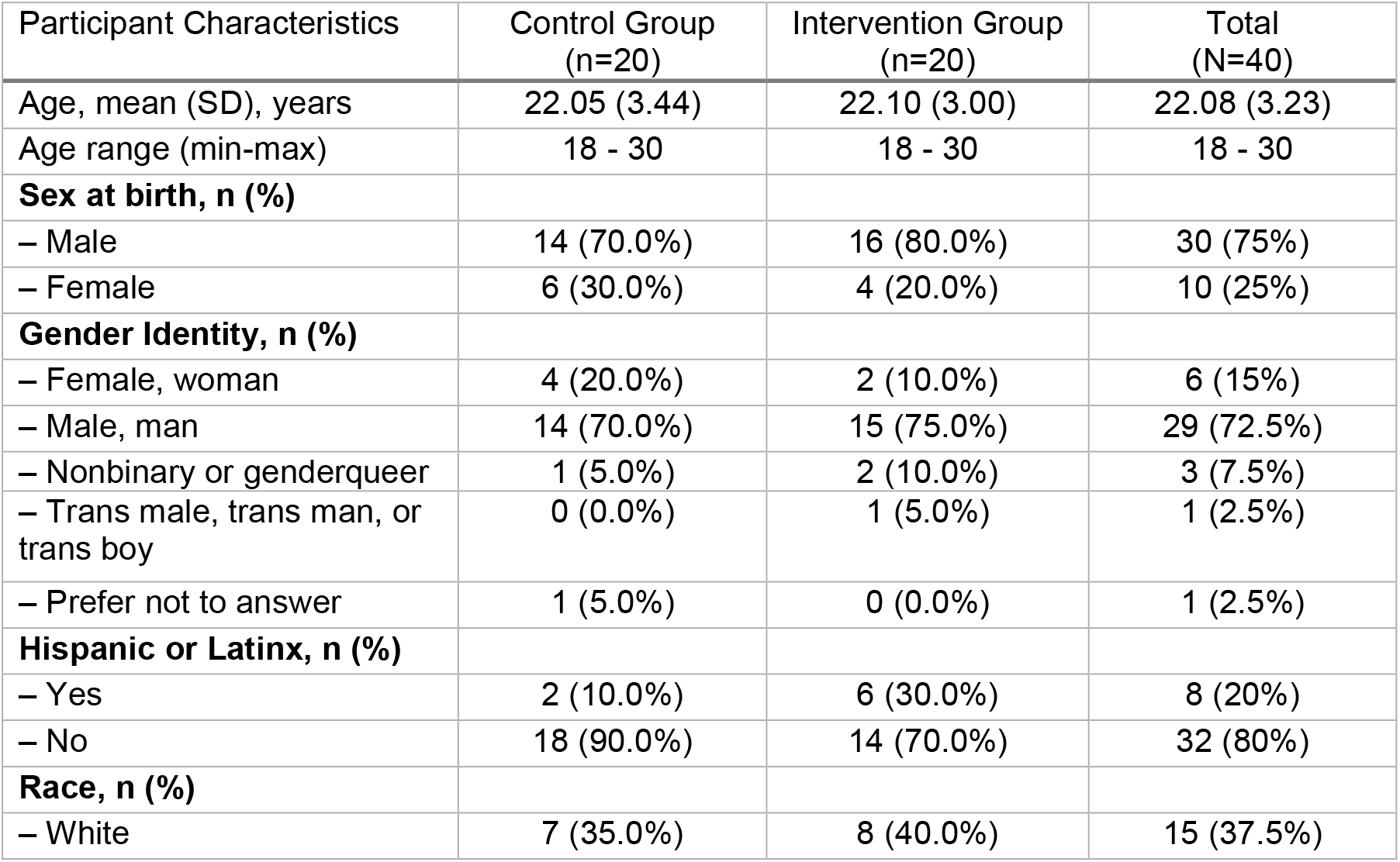
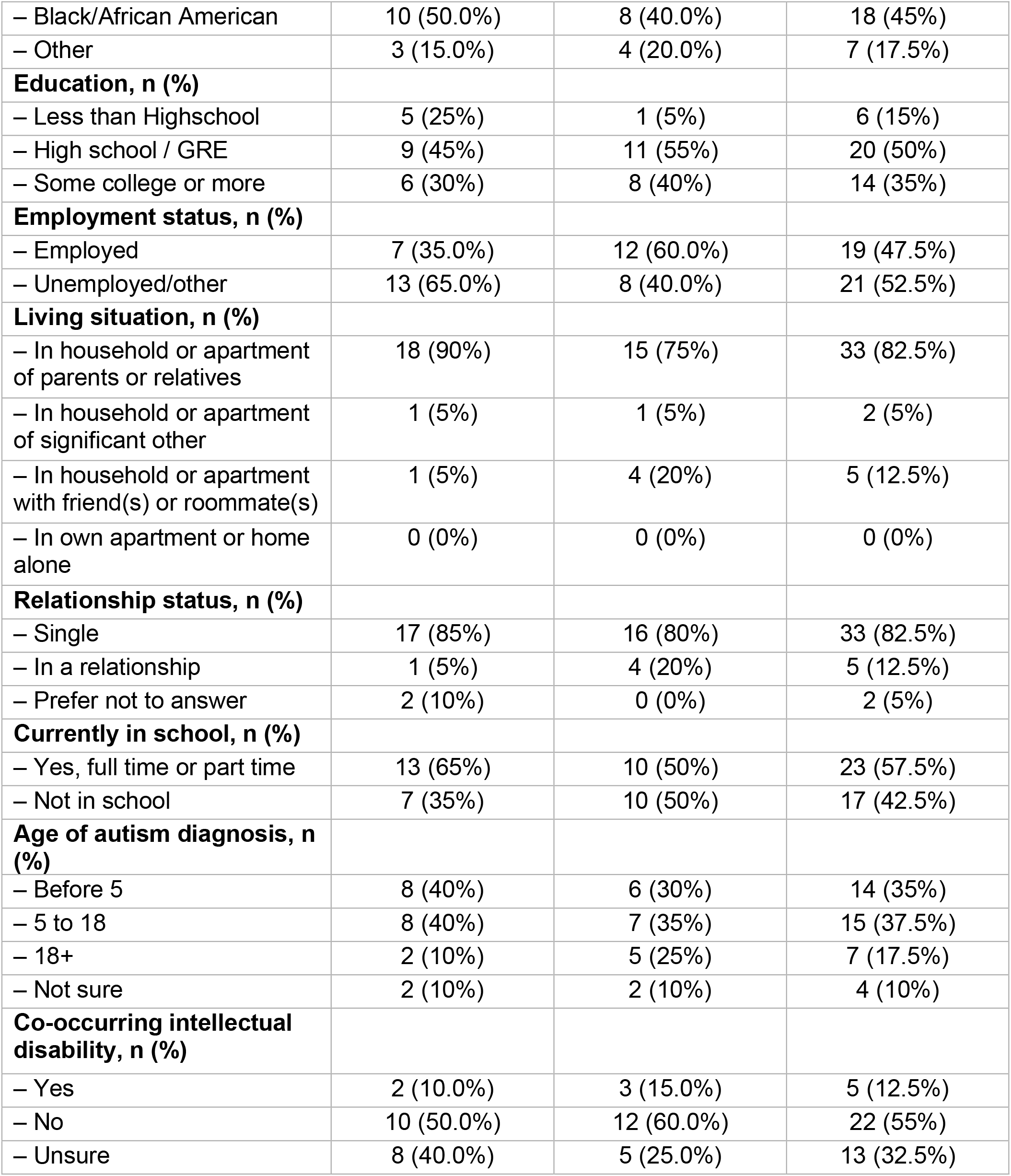
Demographic and clinical characteristics of participants (N=40)

| Participant Characteristics | Control Group<br>(n=20) | Intervention Group<br>(n=20) | Total<br>(N=40) |
| --- | --- | --- | --- |
| Age, mean (SD), years | 22.05 (3.44) | 22.10 (3.00) | 22.08 (3.23) |
| Age range (min-max) | 18 - 30 | 18 - 30 | 18 - 30 |
| **Sex at birth, n (%)** |  |  |  |
| – Male | 14 (70.0%) | 16 (80.0%) | 30 (75%) |
| – Female | 6 (30.0%) | 4 (20.0%) | 10 (25%) |
| **Gender Identity, n (%)** |  |  |  |
| – Female, woman | 4 (20.0%) | 2 (10.0%) | 6 (15%) |
| – Male, man | 14 (70.0%) | 15 (75.0%) | 29 (72.5%) |
| – Nonbinary or genderqueer | 1 (5.0%) | 2 (10.0%) | 3 (7.5%) |
| – Trans male, trans man, or trans boy | 0 (0.0%) | 1 (5.0%) | 1 (2.5%) |
| – Prefer not to answer | 1 (5.0%) | 0 (0.0%) | 1 (2.5%) |
| **Hispanic or Latinx, n (%)** |  |  |  |
| – Yes | 2 (10.0%) | 6 (30.0%) | 8 (20%) |
| – No | 18 (90.0%) | 14 (70.0%) | 32 (80%) |
| **Race, n (%)** |  |  |  |
| – White | 7 (35.0%) | 8 (40.0%) | 15 (37.5%) |
| – Black/African American | 10 (50.0%) | 8 (40.0%) | 18 (45%) |
| – Other | 3 (15.0%) | 4 (20.0%) | 7 (17.5%) |
| **Education, n (%)** |  |  |  |
| – Less than Highschool | 5 (25%) | 1 (5%) | 6 (15%) |
| – High school / GRE | 9 (45%) | 11 (55%) | 20 (50%) |
| – Some college or more | 6 (30%) | 8 (40%) | 14 (35%) |
| **Employment status, n (%)** |  |  |  |
| – Employed | 7 (35.0%) | 12 (60.0%) | 19 (47.5%) |
| – Unemployed/other | 13 (65.0%) | 8 (40.0%) | 21 (52.5%) |
| **Living situation, n (%)** |  |  |  |
| – In household or apartment of parents or relatives | 18 (90%) | 15 (75%) | 33 (82.5%) |
| – In household or apartment of significant other | 1 (5%) | 1 (5%) | 2 (5%) |
| – In household or apartment with friend(s) or roommate(s) | 1 (5%) | 4 (20%) | 5 (12.5%) |
| – In own apartment or home alone | 0 (0%) | 0 (0%) | 0 (0%) |
| **Relationship status, n (%)** |  |  |  |
| – Single | 17 (85%) | 16 (80%) | 33 (82.5%) |
| – In a relationship | 1 (5%) | 4 (20%) | 5 (12.5%) |
| – Prefer not to answer | 2 (10%) | 0 (0%) | 2 (5%) |
| **Currently in school, n (%)** |  |  |  |
| – Yes, full time or part time | 13 (65%) | 10 (50%) | 23 (57.5%) |
| – Not in school | 7 (35%) | 10 (50%) | 17 (42.5%) |
| **Age of autism diagnosis, n (%)** |  |  |  |
| – Before 5 | 8 (40%) | 6 (30%) | 14 (35%) |
| – 5 to 18 | 8 (40%) | 7 (35%) | 15 (37.5%) |
| – 18+ | 2 (10%) | 5 (25%) | 7 (17.5%) |
| – Not sure | 2 (10%) | 2 (10%) | 4 (10%) |
| **Co-occurring intellectual disability, n (%)** |  |  |  |
| – Yes | 2 (10.0%) | 3 (15.0%) | 5 (12.5%) |
| – No | 10 (50.0%) | 12 (60.0%) | 22 (55%) |
| – Unsure | 8 (40.0%) | 5 (25.0%) | 13 (32.5%) |

### High Engagement in the Program

Among the 20 intervention participants, 10 (50%) received CAPS services for more than 300 days and seven received services for between 60 and 300 days. Three participants had minimal or no engagement: one intervention participant discontinued the CAPS services after a week, one had only one contact with the peer specialist, and one participant did not respond to either the peer specialist or the research team after completing the baseline assessment and therefore did not receive any service. On average, intervention participants received services for 258.56 days (SD=123.62, min=1, max=378). CAPS participants had an average of 46 contacts with the peer specialists (SD=35.50, min=0, max=103), averaging 27.93+/-13.71 minutes per contact for an average total of 16.67 contact hours (SD=11.00, min=1, max=37).

### Numerous Indicators of Possible Program Benefit

The primary purpose of the analyses was to identify possible signals of CAPS effectiveness. As expected given the small sample size, results of the GLMM showed no significant group-by-time interaction effects on study outcomes except for overall QOL (F(2, 30.5)=6.10, p=0.019). Adjusted mean QoL increased from 3.50 (SE=0.23) to 3.89 (SE=0.28) in the CAPS condition and decreased from 3.95 (SE=0.23) to 3.65 (SE=0.28) in the control condition over 12 months. Significant time effects were observed in positive identity (F(2, 35.5)=4.65, p=0.016) and self-efficacy (F(2,32,9)=3.75, p=0.034), indicating improvement over time in both groups.

Comparing within-group effect sizes provided additional insight into the pattern and magnitude of change associated with CAPS participation (Table 3). The CAPS condition had consistently larger positive changes on all outcome measures. The largest effect size difference was found on the Overall QoL outcome (d^i^-d^c^=0.60), followed by social QOL (d^i^-d^c^=0.41). Other notable differences favoring the CAPS condition included greater reductions in internalized stigma (ISMI; d=-0.32), needs (CAN; d=-0.26), social responsiveness (SRS; d=-0.26), and loneliness (UCLA Loneliness; d=0.26), and enhanced positive identity (QDIO; d=0.24). The smallest differences in change over time between groups was observed for self-efficacy (d^i^-d^c^=0.05), although results still favored the CAPS condition.

**Table 3.** Means, SDs, and within-group effect sizes (dependent sample Cohen’s d) for baseline and 12-month.

|  | Control (n=17) |  |  | Intervention (n=15) |  |  |  |
| --- | --- | --- | --- | --- | --- | --- | --- |
|  | Baseline | 12-month | Cohen's d | Baseline | 12-month | Cohen's d | Difference (d<sub>i</sub>-d<sub>c</sub>) |
|  | Mean(SD) | Mean(SD) |  | Mean(SD) | Mean(SD) |  |  |
| **Outcomes** |  |  |  |  |  |  |  |
| Internalized stigma | 1.47 (0.51) | 1.35 (0.79) | -0.17 | 1.73 (0.8) | 1.4 (0.51) | -0.49 | -0.32 |
| Self-efficacy | 3 (0.71) | 3.18 (0.73) | 0.25 | 2.73 (0.7) | 2.93 (0.59) | 0.30 | 0.05 |
| Unmet needs | 7.76 (6.69) | 7.47 (5.96) | -0.05 | 6.27 (4.53) | 4.93 (4.11) | -0.31 | -0.26 |
| Social responsiveness | 61.29 (8.65) | 60.76 (9.54) | -0.06 | 64.47 (12.38) | 60.6 (10.32) | -0.32 | -0.26 |
| Positive identity | 29.65 (7.08) | 31.24 (6.2) | 0.23 | 28.73 (4.82) | 30.87 (3.76) | 0.47 | 0.24 |
| Loneliness | 8.35 (3.3) | 7.94 (2.97) | -0.13 | 7.73 (2.37) | 8.07 (2.74) | 0.13 | 0.26 |
| Quality of Life |  |  |  |  |  |  |  |
| Overall QoL | 3.88 (1.05) | 3.59 (1.33) | -0.23 | 3.47 (1.13) | 3.87 (1.06) | 0.37 | 0.60 |
| Social Domain of QoL | 66.24 (25.77) | 65.88 (24.48) | -0.01 | 61.67 (21.74) | 70 (20.51) | 0.39 | 0.41 |

## Discussion

This study assessed whether there are indications of possible positive effects for the Community Autism Peer Support (CAPS) program, a novel, community-based intervention in which trained autistic peer specialists provide individualized support to autistic young adults pursuing self-determined goals related to community living and participation. Because this feasibility trial was not powered to fully assess effectiveness, the findings should be considered exploratory and no definitive claims are made regarding treatment outcomes. This is consistent with the perspective that findings from underpowered studies are at an elevated risk of incorrectly concluding a null effect or that a significant effect of a certain magnitude will be found in a fully powered trial.^32,48^ Nonetheless, across complementary analyses, the consistent pattern and size of results favoring the CAPS condition suggest that the CAPS intervention may offer a promising approach for enhancing the lives of young autistic adults. At the very least, these findings support continued development and rigorous evaluation of autistic-delivered peer support as an approach to meet the community living and participation needs of autistic young adults to place them on a good trajectory for enhanced well-being in the long-term.

One statistically significant effect did emerge for overall quality of life, where participants in the CAPS condition showed greater improvement than the service-as-usual control participants over a period of 12 months. However, this finding would not be retained when accounting for multiple statistical tests. Nonetheless, it is plausible that quality of life may be particularly responsive to the CAPS model because the intervention is individualized around the life domains that participants identify as important. Rather than targeting a single skill or functional outcome, CAPS supports participants in pursuing personally meaningful goals across multiple areas of community life, including relationships, education, employment, leisure and recreation, independent living, and access to community resources. The CAPS program also integrates shared life experiences with individualized action planning, practical problem-solving, and resource navigation to support autistic adults in pursuing personally meaningful goals and increasing community participation.

While other outcomes did not appear to result in a statistically significant treatment effect, nonsignificance in underpowered studies should not be interpreted as conclusive evidence of ineffectiveness.^49^ Consistent with recommendations for small-scale trials, additional emphasis is placed on estimates of change and patterns of outcome rather than statistical significance testing. The CAPS condition demonstrated larger improvements from baseline to 12 months compared to the control condition across all outcomes, providing evidence of a consistent positive signals. Although these results are not definitive, they provide initial support for the importance of pursuing further research on CAPS in large-scale randomized, controlled trials.

Beyond quality of life, the intervention group consistently demonstrated more favorable changes in loneliness, internalized stigma, positive identity, unmet needs, and social functioning, all of which were targeted by the intervention. Taken together, these findings suggest a coherent pattern of improvement across multiple domains that are highly relevant to autistic adults.

Importantly, many of these outcomes have received relatively limited attention in intervention research for autistic adults, despite being identified as important outcomes to target by autistic people ^50–52^. Unlike many existing adult interventions that primarily target specific participation outcomes (e.g., employment, education) or social skills, these outcomes reflect broader aspects of psychological well-being, belongness, and meaningful social roles that are increasingly recognized as important to their lives. The consistent pattern of findings observed in the present study suggests that autistic-delivered services are promising in terms of their possible benefits. Moreover, as a strength-based intervention grounded in shared lived experience, it remains plausible that autistic peer specialists who deliver this type of intervention may complement existing services or interventions by addressing outcomes that have historically been underrepresented in autism intervention research while aligning more closely with priorities identified by autistic adults.

## Limitations

Several limitations are identified. First, it was not feasible to blind the participants to the behavioral intervention owing to its nature. While there was a concern that participants might disengage or drop out once they found out that they had been assigned to treatment as usual, the attrition rate of the control group was low (0% at 3-month, and 15% at 12-month), suggesting that randomization and study procedures did not substantially affect participant retention. Second, the CAPS intervention was intentionally individualized and participant-driven, resulting in substantial variability in intervention dose, duration, and timing across participants. Although this flexibility is consistent with the person-centered nature of peer support, it introduces heterogeneity in intervention exposure and may have reduced the ability to detect treatment effects. Future studies should examine strategies for incorporating analyses that account for intervention dose and timing. Third, as with all randomized trials, randomization does not guarantee equivalence between groups, particularly in small-scale studies. Baseline differences in factors that may influence outcomes, such as autism characteristics, co-occurring mental health conditions, physical health concerns, support needs, and different services received, may have affected the observed results. Larger trials should incorporate sufficient sample sizes to improve balance across groups and consider adjustment for important baseline covariates when appropriate.

## Conclusion

This study provides support for the promise of the CAPS program as an autistic-delivered peer support service for autistic young adults. At a time when relatively few autistic-delivered interventions have been studied, and this intervention focuses on aspects of community living and well-being that are not always well addressed in usual care, CAPS offers a promising, novel service model that combines shared lived experience with individualized, person-centered support. Our data indicated that the program successfully engaged autistic adults from diverse demographics, suggesting broad interest in peer support delivered by autistic adults. Participants remained engaged in CAPS services over extended periods, demonstrating the feasibility of implementing sustained peer support within a community behavioral health setting.

Furthermore, while the study was not powered to establish intervention efficacy, the patterns of findings demonstrated consistent positive signals across multiple participant-centered outcomes, with the strongest improvements observed in the overall and social quality of life. Overall, CAPS demonstrates the potential of leveraging autistic lived experience in the design and delivery of supports that lead to outcomes that are also important and meaningful to the population. These study results provide a foundation for future exploration and testing of CAPS and other similar interventions.

## Acknowledgement

This work was supported in part by Philadelphia’s Department of Behavioral Health and Intellectual disAbility Services and Community Behavioral Health. We thank our community partner organization - Northeast Treatment Centers, for their collaboration in implementing the Community Autism Peer Support (CAPS) program and supporting study activities. In addition, we are deeply grateful to the members of the Community Advisory Board - including autistic adults, family members, self-advocates, and peer specialists, whose guidance, expertise, and lived experiences were instrumental in the development, refinement, and implementation of the CAPS program and this research. Lastly, we would also like to sincerely thank the participants whose support and commitment made this trial possible.

## Authorship Confirmation Statement

W.S: conceptualization, methodology, writing – original draft. A.C.: study design and implementation, writing-review and editing. G.B: implementation, writing-review and editing. K.K.: conceptualization, funding acquisition, and writing-review and editing. J.R.: formal analysis, investigation, writing-review and editing. J.V.: formal analysis, investigation, writing-review and editing. S.N.: study design and implementation, writing-review and editing. L.S.: project administration, funding acquisition, writing-review and editing. M.S.: conceptualization, methodology, writing-review and editing, project administration, and funding acquisition

## Statements and Declarations

### Ethical consideration

This study was approved by the Drexel University IRB (Approval No. 2206009290) and the City of Philadelphia IRB (Approval No.2022-57). All procedures were conducted in accordance with the ethical standards of the institutional research committee and with the 1964 Helsinki Declaration and its later amendments. This manuscript does not contain individual personal data, image, or videos.

### Consent to participate

Electronic consent was obtained from all participants prior to their participation in the study.

### Data availability statement

The datasets generated during and/or analyzed during the current study will be available in the National Institute of Mental Health Data Archive repository.

